# Bacillus Calmette-Guérin (BCG) vaccination: a global analysis of cost-effectiveness and optimal safety stock

**DOI:** 10.64898/2025.12.11.25342124

**Authors:** Debebe Shaweno, Nyashadzaishe Mafirakureva, Andrew Lee, Tuck Seng Wong, Peter. J. Dodd

## Abstract

**Background:** Bacille Calmette-Guérin (BCG) is the most commonly-used vaccination globally, but multi-country cost-effectiveness analyses are outdated and have not considered sequelae in tuberculosis survivors. A rationale is lacking to guide the size of safety stocks.

**Methods:** For the 110 countries using universal neonatal BCG vaccination, we used a decision tree model to compare the costs and health benefits of status-quo BCG vaccination for children aged 0-4 years in 2023 to a counterfactual where no BCG was used, accounting for post-tuberculosis sequelae. We assessed cost-effectiveness against a threshold of 30% per capita gross domestic product from a health system perspective. We determined the safety stock that maximized expected net benefit.

**Findings:** BCG vaccination prevented 742,000 (95% uncertainty interval[UI]: 541,000 to 991,000) tuberculosis episodes and 192,000 (95%UI:138,000 to 264,000) tuberculosis deaths globally. Of these, 49,000 (95%UI: 32,300 to 72,100) episodes and 30,000 (95%UI: 19,600 to 45,100) deaths prevented were from tuberculous meningitis. Universal neonatal BCG vaccination was cost-effective in the majority (75/110) of countries where it is used and in all countries with estimated tuberculosis incidence over 42 per 100,000 per year, with a median incremental cost-effectiveness ratio of $276 (interquartile range: $84 to $1514) per disability-adjusted life-year averted. Median optimal safety stock for a 10% uncertainty in demand was 11% (IQR: 7% to 17%) of expected demand.

**Interpretation:** BCG vaccination continues to prevent substantial morbidity and mortality in children globally, and cost-effectiveness considerations support continued universal vaccination in most countries with this policy currently.

**Funding:** UK EPSRC & DHSC

## Introduction

Tuberculosis is one of the leading causes of morbidity and mortality in children under 5 years of age.^1^ Children are at increased risk of progression to disease following tuberculosis infection, and are more likely than adults to experience severe forms of disease, including tuberculous meningitis (TBM).^2^ The high rates of mortality and neurological sequelae from TBM are estimated to translate into 13,380 deaths and 3,950 children with neurological sequelae in 2019 in this age group.^2^ Bacille Calmette-Guérin (BCG) vaccination offers limited and uncertain protection against pulmonary tuberculosis in adults, but is protective in children and against severe forms of disease. Today, BCG remains one of the world’s most widely used vaccines with over 115 million children receiving BCG across more than 161 countries in 2023.

BCG vaccination is recommended by the World Health Organization (WHO) for all children in high tuberculosis incidence settings as a single dose at birth or selected neonates in high-risk groups in low burden settings.^3^ BCG is relatively cheap, and has been found to be cost-effective in many settings.^4^ In several low tuberculosis incidence settings BCG has been found not cost-effective,^5^ and many countries with low tuberculosis incidence have discontinued universal BCG vaccination in favour of selective vaccination in specific risk groups.

Knowledge and thinking have evolved so as to make re-evaluation of the cost-effectiveness of BCG vaccination timely. Except for the evaluation of Bourdin Trunz and Dye,^4^ which only considered TBM, previous economic evaluations have not included TBM. We now have a stronger understanding of the proportion of tuberculosis in children that is TBM, which is expected to be a major driver of cost and health impacts. There is also a better understanding of long-term post-tuberculosis sequelae, and a movement towards including these increases in mortality and reductions in quality of life in economic evaluations of preventive interventions.^6^ At the same time, there is now recognition that previous guidance suggested interventions could be considered cost-effective with incremental cost-effectiveness thresholds (ICERs) per disability-adjusted life-year (DALY) averted below 1-3 x per capita gross domestic product (GDP) was misleading. Newer analyses suggest the lowest ICERs for components of health systems are typically in the range 0.3 - 0.5 x GDP, representing a much stricter criterion for cost-effectiveness.^7^

Despite its global importance, BCG supply has been subject to disruptions in recent years due to limited supplier options and flexibility - only four of 25 existing suppliers are WHO-prequalified and used by UNICEF, with one supplier accounting for 70% of production capacity. These disruptions have resulted in stock-outs, which have the potential to cause considerable increases in mortality.^8^ Often a safety stock buffer of 25% is used,^9^ but this recommendation is not evidence-based. Health economics provides a rational utility-maximizing framework for determining optimal safety stocks in the face of uncertain demand, which requires economic evaluation of BCG vaccination at a country level.

We therefore re-evaluated the global cost-effectiveness of BCG vaccination against various choices of cost-effectiveness threshold, accounting for TBM and post-tuberculosis sequelae. We also developed a method to determine optimal BCG safety stocks, and calculated these for various levels of uncertainty in demand.

## Methods

### Optimal safety stock

For a given cost-effectiveness threshold, the expected net benefit (ENB) captures the utility associated with a strategy incorporating health gains in monetary units less the economic cost of the intervention. Figure 1A shows the expected dependence of ENB on number of vaccine doses for a fixed demand *D*: where BCG use is cost-effective, we gain ENB at a rate of *g* per dose procured and used; once volume exceeds demand, additional doses subtract from ENB at the rate of *h* per dose procured. In general, the demand has uncertainty quantified by a probability distribution with quantile function *Q*. In the appendix, we show that the optimal vaccine volume maximizing ENB for an uncertain demand is given by *V* = *Q*(*g*/(*g* + *h*)). To calculate *g* and *h* we undertake a cost-effectiveness analysis of BCG use, extracting the expected gain in ENB per dose used (*g*), and the expected decrease in ENB per dose wasted (*h*).

**Figure 1.**
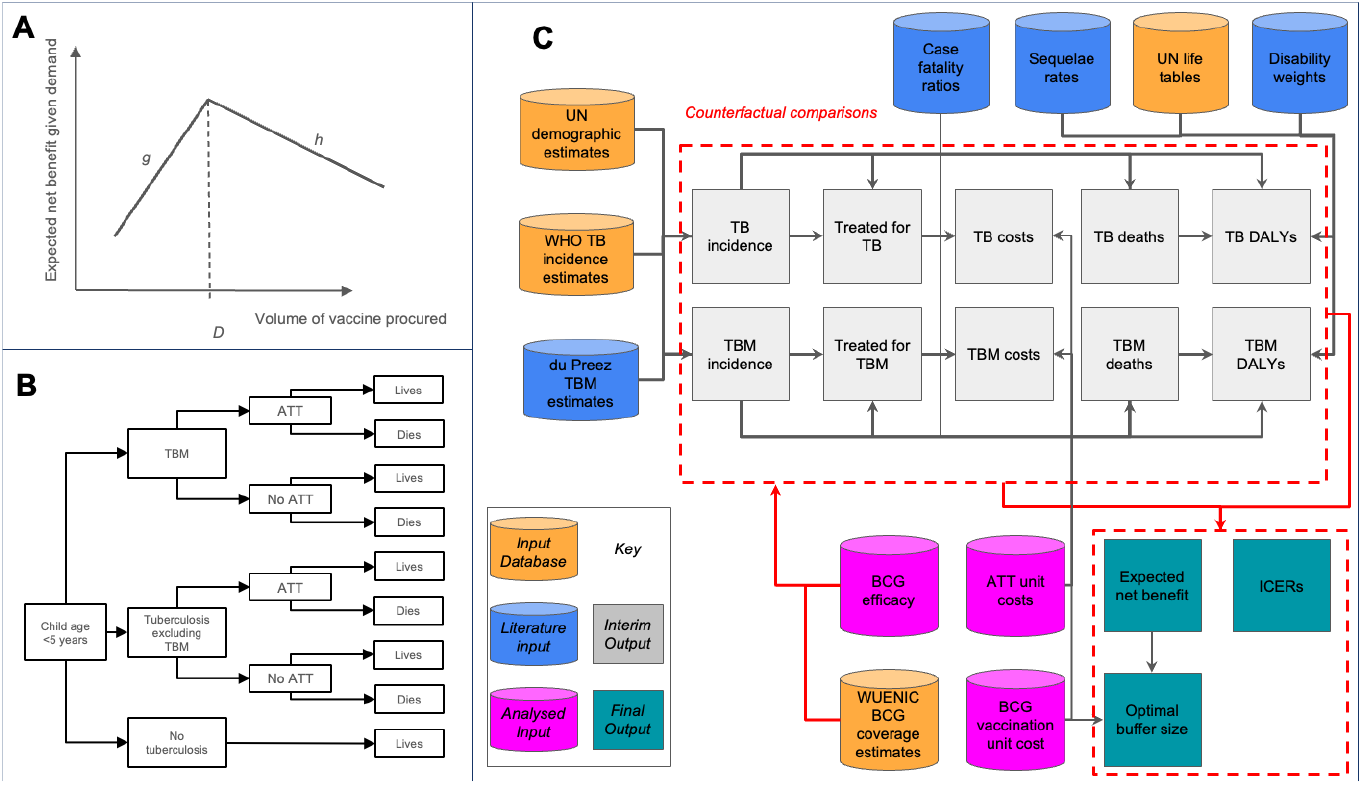
Representation of methods used. **A** Expected net benefit as a function of vaccine procured for known demand, *D*. **B** Structure of decision tree model used in economic evaluation. **C** Schematic workflow including data inputs and output metrics. TBM = tuberculous meningitis; ATT = anti-tuberculosis treatment; UN = United Nations; DALY = disability-adjusted life-year; TB = tuberculosis; BCG = Bacillus Calmette-Guérin; WUENIC = WHO/UNICEF estimates of immunization coverage; ICER = incremental cost-effectiveness ratio.

### Health economic approach

We undertook a cost-utility analysis from a health systems perspective at country-level, comparing status quo BCG vaccination use in children aged 0-4 years in 2023 with a counterfactual where no BCG was used. We used a life-time horizon for benefits and assumed that costs accrued in the present. We used a 3% discount rate in line with the iDSI reference case.^10^ Outcomes were measured in disability-adjusted life years (DALYs) including life-years lost due to deaths from tuberculosis, and for our base case, including post-tuberculosis increases in mortality rates and decreases in quality of life. Sensitivity analyses included omitting post-tuberulosis morbidity and mortality effects.

A decision tree model (Figure 1B) was used to compute outcomes and costs, and for our base case included TBM. Probability of death and post-tuberculosis morbidity depended on whether tuberculosis was TBM, and whether children received anti-tuberculosis treatment (ATT). Sensitivity analyses included omitting TBM from the model. BCG vaccination was assumed to reduce the risk of incident tuberculosis disease, and the probability that tuberculosis disease is TBM should it occur (see next section). Full details of the model and its parametrization are in the Appendix. Data sources and analysis workflow are shown schematically in Figure 1C. We undertook our analysis for all countries appearing in WHO/UNICEF Estimates of National Immunization Coverage (WUENIC) BCG coverage estimates.

We included resources used in purchasing, administering and delivering BCG vaccination doses, basing unit costs on country-specific standardised estimates generated from the Immunisation Delivery Cost-Coverage (IDCC) database.^11^ We also included calculated unit costs of ATT for TBM and tuberculosis excluding TBM following an approach we previously described in Dodd and colleagues.^12^ Costs were calculated as the sum of outpatient care costs, inpatient care costs, national tuberculosis programme costs, and the costs of drugs based on country reported data in the WHO tuberculosis database. We accounted for additional resource use due to TBM by inflating inpatient care costs using an adjustment factor derived from Peng et al.^13^ and assuming first-line anti-tuberculosis drugs are given for twice the standard duration. Costs were adjusted for inflation to 2023 US dollars ($) using inflation rates from the World Bank.

The health impacts on tuberculosis survivors included increased mortality rates and reductions in health-related quality of life that were larger in the first three years following tuberculosis disease.^14^ To capture reductions in health-related quality of life following TBM, we did a meta-analysis of existing systematic review data to determine the fraction of TBM survivors with severe sequelae, to which we applied a disability weighting based on severe motor and cognitive impairments GBD 2013^15^ (see Appendix).

To capture uncertainty, probabilistic sensitivity analysis was used. All model data and parameter inputs were treated as uncertain, and 10,000 samples from the distributions described in the Appendix were used to evaluate outcomes for each country.

We did not write a health economic analysis plan, engage those affected by tuberculosis in designing the study, or include heterogeneity or subpopulations.

### BCG efficacy

We took the efficacy of BCG in preventing incident tuberculosis from the estimate in the systematic review and meta-analysis of Martinez et al.^16^ as 0.63 (0.49 - 0.81). Our model required the efficacy of BCG in preventing TBM conditional on tuberculosis disease, which does not seem to have been previously reported. In order to calculate this, we re-analysed a subset of data presented in Abubakar et al.^17^ with tuberculosis stratified as TBM or not. We used Mantel–Haenszel (MH) random effects meta-analysis fitted via restricted maximum likelihood to pool risk ratios of TBM (protection against TBM conditional on having TB disease, i.e. using TBM as a numerator from incident tuberculosis as a denominator in a binomial likelihood).

### Metrics reported

Globally, and for each WHO region, we calculated the status quo, counterfactual (no BCG vaccination), and incremental resource use, economic costs, and health outcomes for 2023. Resource use is quantified in terms of BCG vaccinations, ATT courses, and hospitalizations; economic costs are reported associated with each of these resources and in total; health outcomes are reported in terms of TBM and tuberculosis incidence, deaths, sequelae and DALYs. We also calculate the total and per capita tuberculosis deaths averted by status quo vaccination for each country, and resource use, costs and health outcomes for the top 10 countries by averted tuberculosis deaths. We calculate incremental cost-effectiveness ratios (ICERs) for each country and compare these against indicative cost-effectiveness thresholds of 0.3x, 0.5x, and 1x GDP, and report them as a ratio of GDP. We report uncertainty in cost-effectiveness in the form of quantiles for cost-effectiveness acceptability curves in each country, and explore the determinants of cost-effectiveness through regression analysis. Finally, we also calculate optimal buffer size as a percentage of expected demand, assuming normally distributed uncertainty in demand with coefficients of variation equal to 5%, 10%, and 15%.

### Role of the funding source

The funder of the study had no role in study design, data collection, data analysis, data interpretation, writing of the manuscript or decision to submit.

## Results

Our analysis of the conditional effect of reducing the risk of TBM in tuberculosis included six studies (see figure 2a), with 4,617 people aged 0-14 years with tuberculosis, 50.6% of whom had BCG vaccination and 3.4% of whom had TBM. Random-effects meta-analysis yielded a pooled estimate on the RR of TBM conditional on TB of 0.58 (95% confidence interval, 0.20, 1.74) with heterogeneity of *I*^*2*^ = 71.6%. Our analysis of TBM sequelae found 56% of sequelae were categorized as severe (see Figure 2b), resulting in a mean disability weighting among all TBM survivors of 0.34.

**Figure 2.**
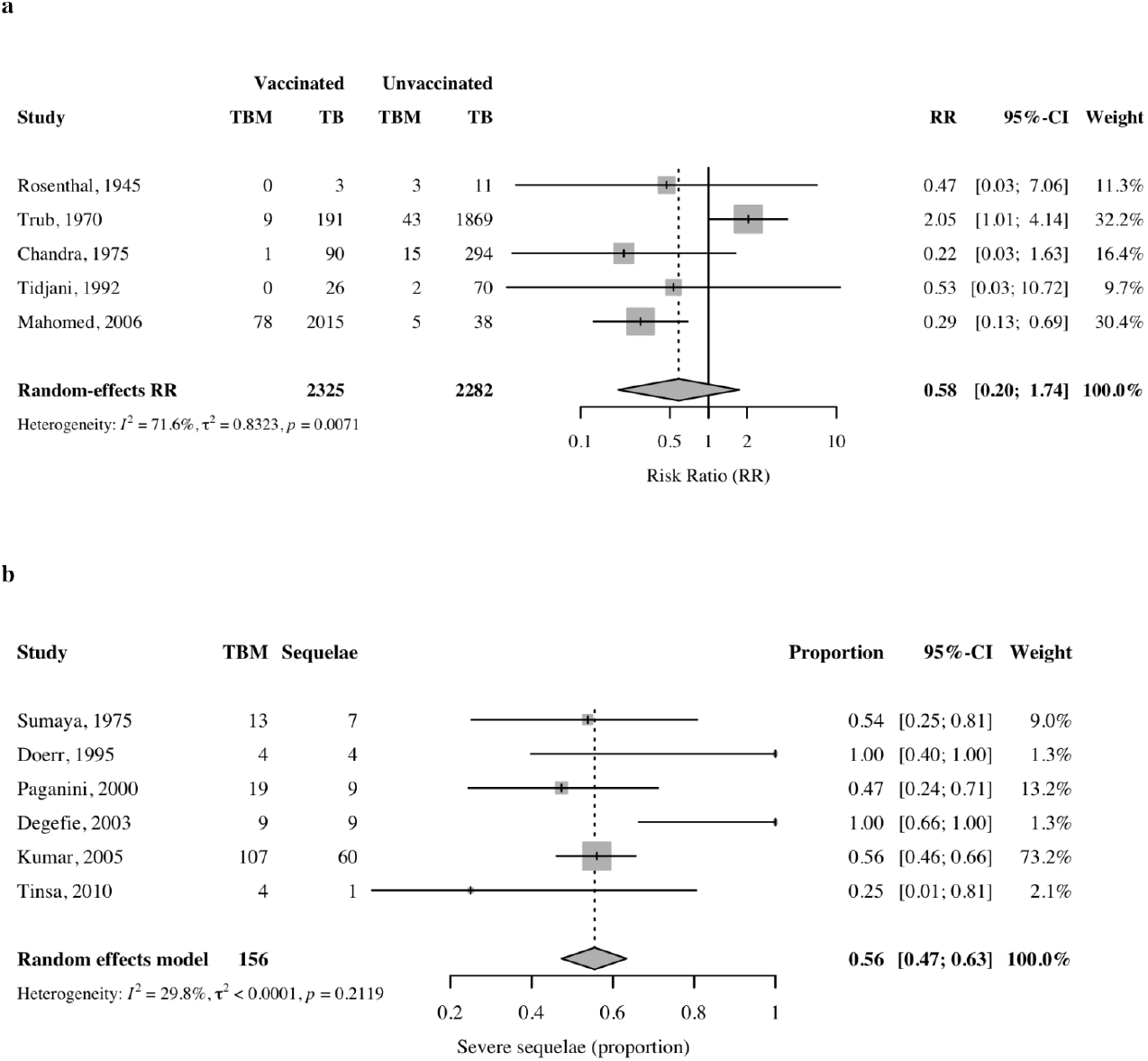
Forest plots from meta-analyses. **a)** Efficacy of BCG vaccination in preventing tuberculous meningitis in children under 5 years of age who have developed tuberculosis disease **b)** Proportion of sequelae classed as severe among TBM survivors. Data from Abubakar et al.^17^ for a) and Chiang et al.^32^ for b)

Our modelling analysis of BCG included 110 countries, and a total of 558 million children under 5 years of age. This represents 95% of the global population in this age group, and 99% of WHO-estimated global tuberculosis incidence in this age group.

The median unit cost of vaccine delivery per BCG dose (appendix figure S4), adjusted to 2023 prices, was $3.1 (95% IQR:1.8-4.9). The median cost per ATT (appendix figure S3) was $576 (319 - 1455) for children without TBM and $618 (344 - 1678) for children with TBM.

Under the status quo scenario, 496 million of these children received BCG vaccination (Table 1), equating to an aggregate coverage of 87%, and ranging from 37% to 99% in individual countries. The total economic cost of utilized BCG vaccination was estimated at $1,070 (845 to 1,350) million.

**Table 1.** Changes in total resource use, costs and health outcomes globally for children aged <5 years in 2023.

| Item | Status quo | Counterfactual (no BCG) | Counterfactual (no BCG) - Status quo |
| --- | --- | --- | --- |
| **Resource** |  |  |  |
| BCG doses | 496 (496 to 496) M | 0 (0 to 0) M | -496 (-496 to -496) M |
| ATT for TB* | 277,000 (216,000 to 339,000) | 636,000 (479,000 to 815,000) | 359,000 (254,000 to 492,000) |
| TBM hospitalizations | 9,390 (6,690 to 12,700) | 33,000 (22,100 to 48,000) | 23,600 (15,000 to 36,000) |
| **Cost(\$M)** | | | |
| BCG vaccination cost | 1,070 (845 to 1,350) | 0 (0 to 0) | -1,070 (-1,350 to -845) |
| ATT cost for TB* | 84 (66 to 103) | 194 (149 to 246) | 110 (81 to 147) |
| ATT cost for TBM | 3.2 (2.4 to 4.4) | 11 (8 to 16) | 8.3 (5.5 to 12) |
| All costs | 1,120 (898 to 1,410) | 194 (149 to 246) | -927 (-1,220 to -701) |
| **Health** |  |  |  |
| TB* incidence | 569,000 (452,000 to 688,000) | 1,310,000 (1,010,000 to 1,650,000) | 742,000 (541,000 to 991,000) |
| TBM incidence | 19,300 (14,100 to 25,600) | 68,400 (47,400 to 96,200) | 49,000 (32,300 to 72,100) |
| TB* deaths | 140,000 (108,000 to 173,000) | 333,000 (252,000 to 429,000) | 192,000 (138,000 to 264,000) |
| TBM deaths | 11,700 (8,460 to 15,900) | 41,700 (28,700 to 59,800) | 30,000 (19,600 to 45,100) |
| TBM severe sequelae | 4,240 (3,020 to 5,760) | 14,900 (9,950 to 21,900) | 10,600 (6,720 to 16,400) |
| DALYs from TB* | 7.8 (6.2 to 9.5) M | 18 (14 to 23) M | 10 (7.2 to 13) M |
| DALYs from TBM | 0.094 (0.067 to 0.13) M | 0.33 (0.22 to 0.48) M | 0.24 (0.15 to 0.36) M |

*BCG =Bacillus Calmette-Guérin, TB=Tuberculosis, TBM=Tuberculosis meningitis, ATT= Anti-TB treatment, DALYs= Disability Adjusted Life Years, M = Million*

In the counterfactual scenario with no BCG vaccination, tuberculosis incidence would increase by a total of 742,000 (541,000 to 991,00) in this group (TBM incidence by 49,000 (32,300 to 72,100)). This increase would generate 192,000 (138,000 to 264,000) additional tuberculosis deaths (30,000 (19,600 to 45,100) from TBM), and an additional 10 million DALYs from tuberculosis (240,000 from TBM). The additional 359,000 ATT courses and 23,600 hospitalizations for TBM were estimated to generate an increase in costs for these activities of $927 million. In summary, 119 new tuberculosis episodes (8 TBM), 28 tuberculosis deaths (5 TBM deaths), and 1653 tuberculosis DALYs (45 TBM DALYs) were averted per 100,000 BCG doses.

These impacts varied by setting, with the largest impact in the WHO South-East Asia region (appendix Figure S8). The top 10 countries by tuberculosis deaths averted were India, Indonesia, China, Pakistan, Bangladesh, Myanmar, Nigeria, Philippines, Angola and Tanzania (see appendix table S2). Restricting to countries with > 1,000 tuberculosis notifications in 2023, reductions in per capita tuberculosis mortality and incidence were greatest in Mongolia (150 deaths averted per 100,000 BCG doses), and Indonesia (740 episodes averted per 100,000 BCG doses), respectively.

Compared against a benchmark cost-effectiveness threshold of 0.3 x GDP, BCG was cost-effective in the majority (68%, 75 from 110) of countries where it is used (see figure 3). The median ICER was $276 (interquartile range [IQR]:84 to 1514) per DALY averted. The median ICER/GDP ratio was 0.11(IQR 0.03 to 0.54) see appendix Figure S9). Uncertainty in cost-effectiveness, as measured by cost-effectiveness acceptability curve quantiles, was large (median of IQR/median = 1.26) and varied by region (see appendix figure), with the WHO WPR region having the highest proportional spread (median of IQR/median = 1.38), followed by the WHO Africa region (median of IQR/median = 1.34. Regression analysis of the ENB showed that per capita tuberculosis incidence was a key determinant of cost-effectiveness, explaining 91% of the variance. BCG vaccination was cost-effective at a threshold of 0.3 x GDP for all countries with per capita tuberculosis incidence above 41 per 100,000 person-years. It was not cost-effective in all countries where the incidence is less than 7 per 100,000 (appendix Figure S11)

**Figure 3.**
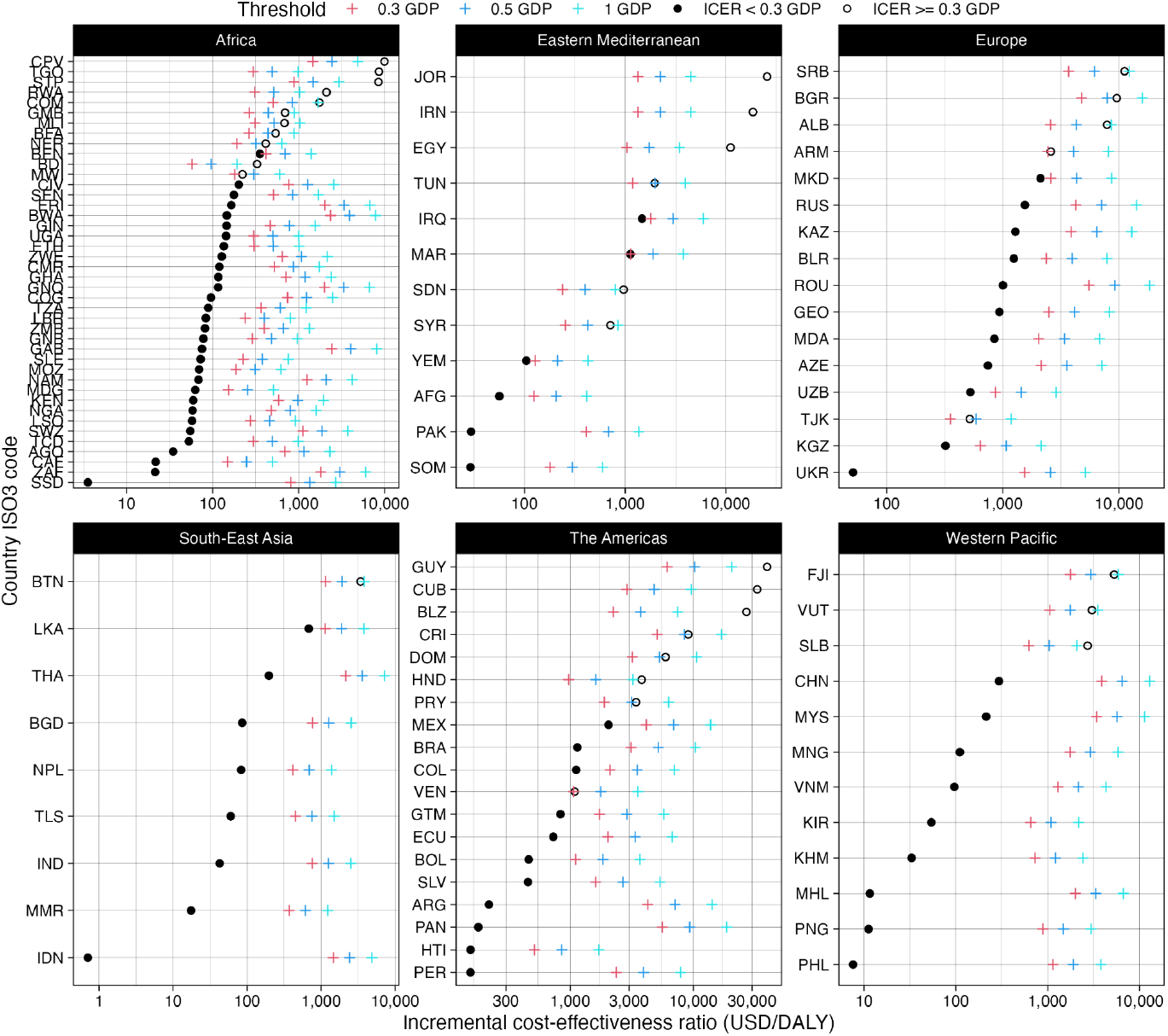
Incremental cost-effectiveness ratios of BCG vaccination in children age <5 years in 2023 compared with no BCG vaccination. ICER = Incremental cost-effectiveness ratio; GDP = (per capita) gross domestic product; USD = United States dollar; DALY = disability-adjusted life-year. Dots represent ICERs and are solid when considered cost-effective against a benchmark of 0.3x GDP. Note the log scale.

Sensitivity analyses omitting post-tuberculosis morbidity and TBM worsened cost-effectiveness, with 64% of countries being cost-effective at a threshold of 0.3 x GDP, and the median ICER changing to $449 (IQR:123 to 2536). Omitting post-tuberculosis effects had a higher effect on cost-effectiveness, with 65% of countries being cost-effective at a threshold of 0.3 x GDP, and the median ICER changing to $393 (IQR:114 to 2211). Omitting TBM had a smaller effect, with 68% of countries being cost-effective at a threshold of 0.3 x GDP, and the median ICER changing to $292 (IQR:87 to 1594). Changes under these sensitivity analyses varied by region, with larger increases in ICERs for the WHO Americas, Europe, and Western Pacific regions (see appendix Figure S12).

For a cost-effectiveness threshold of 0.3x GDP and a proportional uncertainty in demand of 10%, the median optimal safety stock for countries where BCG was cost-effective was 11.6% (IQR 6.7 to 16.9); see Figure 4). For larger uncertainty (CV=15%) this rose to 17.2% (IQR 10 to 25.4) and for smaller uncertainty (CV=5%) it fell to 5.7% (IQR =3.4 to 8.5). The level varied by region, with the highest optimal safety stocks in the Western Pacific region, 16.9% (IQR =16.8 to 19.7). Higher cost-effectiveness thresholds would result in larger optimal safety stocks.

**Figure 4.**
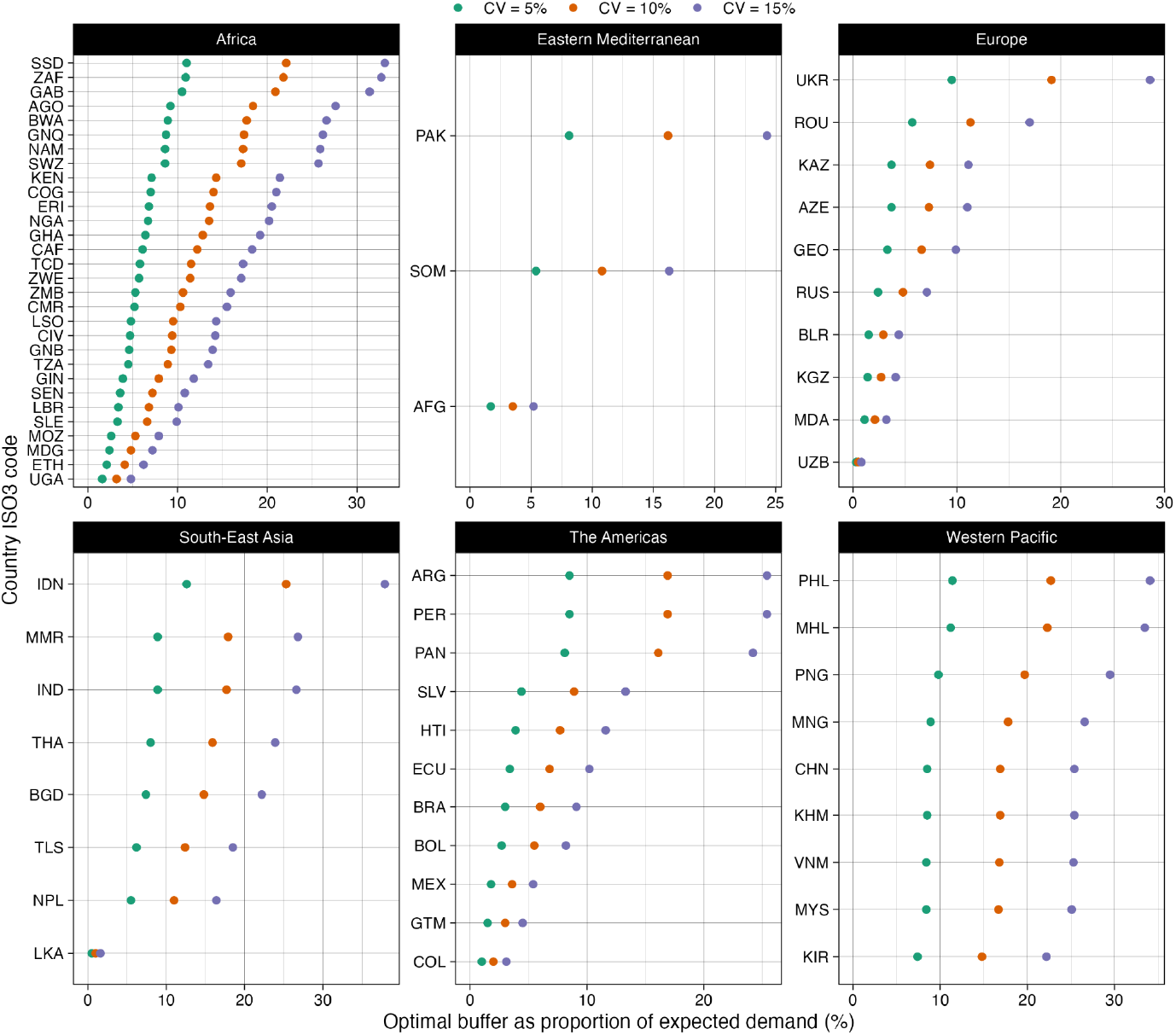
Optimal safety stock as a proportion of expected demand for a cost-effectiveness threshold of 0.3 x GDP and different demand uncertainty. GDP = (per capita) gross domestic product; CV = coefficient of variation for distribution representing demand uncertainty.

## Discussion

We found that levels of BCG vaccination in 2023 made substantial contributions to preventing tuberculosis burden in children aged 0-4 years globally, averting 742,000 tuberculosis episodes and 192,000 tuberculosis deaths. These levels of BCG vaccination also averted 49,000 TBM episodes, which were associated with 30,000 deaths and severe outcomes in 10,600 TBM survivors. Including sequelae for all forms of tuberculosis, we found BCG vaccination in 2023 averted 10 million DALYs.

Few analyses have considered the cost-effectiveness of BCG across multiple countries.^5^ Most economic evaluations have focused on high income, low tuberculosis incidence settings,^18–22^ have focussed on strategies to improve technical efficiency,^23,24^ BCG revaccination,^25,26^ or have not valued benefits in DALYs.^27^ Only Bourdin Trunz and Dye^4^ in 2006 have performed a multi-country cost-utility analysis of global scope, and they included only TBM and disseminated tuberculosis as outcomes, used a single cost per BCG dose based on expert opinion, and did not account for averted tuberculosis treatment costs or post-tuberculosis reductions in health-related quality of life.

Our health economic analysis estimated a median ICER of $276 (IQR:84 to 1514) per DALY averted, which is comparable with other interventions that are considered cost-effective pillars of health systems.^28^ This is also comparable with the global ICER of $206 per DALY averted found by Bourdin Trunz and Dye.^4^ The choice of thresholds below which to consider interventions cost-effective is ultimately a matter for country decision-makers. However, analyses of health expenditure have led to withdrawal of guidance to consider 1-3x per capita GDP a threshold for cost-effectiveness, as in most countries the least cost-effective component of the health system has an ICER of 0.3-0.5 x GDP.^7^ We found that BCG vaccination was cost-effective against a threshold of 30% of per capita GDP in the majority of countries where it is used. Estimated tuberculosis incidence rate was the most important determinant of cost-effectiveness: we found BCG was cost-effective in all countries with estimated tuberculosis incidence rates of over 42 per 100,000 per year, and for no countries with tuberculosis incidence rates of under 7 per 100,000 per year. In the intermediate range, BCG was still cost-effective in the majority of countries where it is currently used.

In 1994, the International Union Against Tuberculosis and Lung Disease set out consensus criteria for the discontinuation of BCG vaccination in countries with low tuberculosis prevalence.^29^ These were defined in terms of smear-positive tuberculosis notification rates being below 5 per 100,000 per year, TBM rates in children being under 1 per million per year, and annual risks of tuberculosis infection being below 0.1%. The most recent WHO position paper re-affirms the use of universal BCG vaccination in countries with tuberculosis incidence rates above 40 per 100,000 per year.^3^ This threshold tallies very closely with the level of incidence above which we found universal BCG to be cost-effective in all countries analysed. However, cost-effectiveness is not the only criteria for switching from universal to selective BCG vaccination, and while encouraging reassessment in settings with declining tuberculosis rates, the WHO position emphasizes the importance of strong surveillance systems for pulmonary tuberculosis and TBM. In many settings with low average tuberculosis incidence, BCG vaccination may continue to be desirable in key groups or areas.

Disruptions to global BCG supply have resulted in shortages,^30,31^ and there is still demand uncertainty due to demographic and behavioural factors. A rule of thumb of 25% for safety stock has been used, but there does not seem to have been previous analysis to justify safety stock margins. Our novel analysis suggests that for proportional uncertainty in expected demand in the region of 10%, buffers in the region of 12% would maximize expected net benefit, but with non-negligible chance of shortfalls.

We found no published data on uncertainty in country demand estimates, but this uncertainty could be estimated by countries from historic data on fluctuation in demand or demand/uptake differences, or by expert elicitation. While our main analysis assumed low proportional uncertainty around expected demand, described by a Gaussian distribution, the general result encompasses more general distributions that could include low-probability, high-consequence events. Our approach to determining optimal vaccine safety stocks within a health economic utilitarian framework could be applied to other vaccines or pathogens.

Several analyses have considered local operational research approaches to reducing wastage and increasing coverage,^23,24^ but work on understanding improvements to supply chain configurations locally and internationally is lacking. To mitigate the risks associated with concentrated supply, where a small number of manufacturers provide most of the globally available BCG, several global and regional strategies are being advanced. These include efforts to diversify production, such as the Partnerships for African Vaccine Manufacturing (PAVM) and the ASEAN Vaccine Security and Self-Reliance (AVSSR) initiative, as well as expansions of existing manufacturing capacity by major producers (e.g., Merck). Additional measures focus on enhancing supply-chain resilience through long-term procurement agreements with WHO-prequalified suppliers, multi-sourcing of critical raw materials, and strengthening partnerships and technology-transfer mechanisms to support sustainable, geographically distributed vaccine production.

Our analysis has a number of limitations. We used single efficacies of BCG vaccination against tuberculosis disease and in reducing the fraction of tuberculosis episodes that are TBM in children aged under 5 years. While these parameters were treated as uncertain, we did not incorporate potential variation in efficacy by country or by age. We assumed no BCG efficacy beyond the age of 5 years, no non-specific mortality benefits from BCG vaccination, and no effects reducing transmission. Neglecting each of these may have underestimated health benefits from BCG vaccination. We also did not consider benefits from protection against leprosy, Buruli ulcer, or other non-tuberculous mycobacteria. Our estimates of unit costs made use of standard regression approaches to impute treatment costs for countries where these were missing, and we did not consider economies of scale, or drug-resistant tuberculosis. Finally, our framework for optimal safety stock assumed that orders in excess of demand would be wasted, and did not allow for resale or long-term storage.

Particular strengths of our analysis included explicit incorporation of TBM alongside other forms of tuberculosis disease, and accounting for post-tuberculosis reductions in health-related quality of life among tuberculosis survivors, which is increasingly recognized as an important benefit of tuberculosis prevention. TBM is the most devastating form of tuberculosis, and in our analysis we found 56% of sequelae among TBM survivors were classified as severe, which included quadriparesis, severe intellectual impairment, blindness, deafness or inability to move without support. Evidence quantifying quality of life among paediatric tuberculosis survivors is still emerging, and we treated severe TBM sequelae as similar to severe motor and cognitive impairments reported in GBD 2013 disability weightings.^15^ We found that omitting both components of our analysis (TBM and post TB) decreased expected net benefit by a median factor of around 63%, but that BCG remained cost-effective at a threshold of 30% per capita GDP for 64% of countries.

In conclusion, BCG vaccination remains cost-effective in the vast majority of countries where it is used, even when compared against more stringent revised benchmarks. We found BCG vaccination was cost-effective in all countries with tuberculosis incidence greater than 42 per 100,000 per year, and continues to avert a very substantial burden of disease and disability. Where proportional uncertainty in demand is 10%, a safety stock of approximately 12% of expected total demand is optimal in terms of cost-utility for many settings.

## Data Availability

All code and data to reproduce this analysis is publicly available on GitHub at https://github.com/Debebe/Bbuff

## Contributions

### Conceptualisation

PJD. Data collection: DAS. Formal analysis: DAS, PJD, NM. Writing - original draft: PJD, DAS. Writing - review & editing: all authors.

## Declarations of interest

None declared.

## Acknowledgements

PJD, DSA, AL and TSW acknowledge research funding through The UK-South East Asia Vaccine Manufacturing Research Hub. This research is funded by the Department of Health and Social Care using UK International Development funding and is managed by the EPSRC. The views expressed in this publication are those of the authors and not necessarily those of the Department of Health and Social Care.

## Notes

### Competing Interest Statement

The authors have declared no competing interest.

